# Influences on limited antimicrobial use in small-scale freshwater aquaculture farms in central Thailand

**DOI:** 10.1101/2024.02.11.24302655

**Authors:** Nour Alhusein, Boonrat Chantong, Sarin Suwanpakdee, Anuwat Wiratsudakul, Virginia C. Gould, Kantima Wichuwaranan, Kornrawan Poonsawad, Varapon Montrivade, Nutcha Charoenboon, Luechai Sringernyuang, Matthew B. Avison, Helen Lambert, Walasinee Sakcamduang, Henry Buller, Kristen K. Reyher

## Abstract

Recent years have shown substantial growth both in the scale and the spread of freshwater aquaculture in Thailand, raising concerns about potential widespread antimicrobial use. This mixed-methods study used surveys and qualitative interviews to examine conditions of freshwater aquaculture farming in central Thailand in relation to animal health, disease management and patterns of antimicrobial use. Freshwater aquaculture in this area of Thailand was largely a domestic venture operated as a source of additional household income to increase financial security. Aquaculture was often integrated with other types of farming; initial outlay was reduced by repurposing unused crops, food, or animal manure (e.g. chicken droppings and pig dung) to fertilise aquaculture ponds. Among farmers representing twenty farms who were surveyed during 2019, only six farmers said they used antimicrobials. These included oxytetracycline, enrofloxacin, norfloxacin, ciprofloxacin and sulphonamides. Farmers doubted the benefits of using antimicrobials to treat aquatic animals; some believed antimicrobials stunted growth. The high cost of medicines and prohibitive regulations also discouraged antimicrobial use. Farmers linked disease occurrence to changes in the weather, the emergence of new diseases and variable water quality. They relied on farm management practices to maintain the health of their aquatic animals, using lime and salt to maintain and improve water quality and pH and to disinfect aquaculture pools. Farmers also reported obtaining juvenile fish and shrimp selectively from farms known to produce healthy stock. Specialised veterinary services for aquatic farming were rare, so farmers relied on their own experimentation with medicines, peer advice and recommendations of shopkeepers who sold both aquatic feed and medicines. This study unexpectedly reveals limited use of antimicrobials linked to socio-economic and ecological features of small-scale family aquaculture farms.

## Introduction

Fast-developing aquaculture sectors in several Southeast Asian countries have been seen as contributors to the rapidly expanding and diversifying antimicrobial resistance (AMR) documented in animal production systems over the last two decades [1–3]. Analysis of point prevalence surveys reporting AMR from aquatic food animals in Asia has found concerning levels of resistance to medically important antimicrobials in foodborne pathogens [4]. Increased antimicrobial use (AMU) is associated with the emergence of AMR in bacteria, with significant implications for both animal and human health [5]. Schar et al. estimated that by 2030, AMU in aquaculture will constitute 5.7% of global AMU and will carry the highest use intensity per kilogram of biomass [6].

Thailand has been part of the Asian aquaculture surge, being ranked 13^th^ among aquaculture country producers with a production of 889,891 tonnes of live weight in 2017 [2]. Its industries predominantly centre on coastal production of shrimp, molluscs and prawn in salt-water ponds. More recent years, however, have seen the significant growth of inland, fresh-water aquaculture in Thailand [7]. Here the early emphasis has been rather more on the production of fish species, notably catfish and tilapia, though shrimp production has grown rapidly and displayed a similar dynamism to that of the coastal regions [8]. In 2015, freshwater aquaculture in Thailand accounted for 45% of total aquaculture production value, with the remainder from marine/coastal aquaculture [7].

The growth of inland Thai aquaculture (both shrimp and fish) has given rise to many debates, despite its very clear profitability for those involved [9,10]. There are concerns about the sustainability of the older model of domestic rice production in many areas of central Thailand - the widescale and highly profitable conversion of paddy fields to aquaculture ponds has accompanied this transition, changing the nature and regulation of the rural communities that rice traditionally sustained [8,11,12]. A second concern has been the use, over-use and management of fresh-water resources, both river and canals, as critical and replenishable growing media for aquatic species at a time when other uses of water are being employed, particularly as a means of moving effluent and wastewater out of sites of production. Little empirical evidence currently exists as to whether inland aquaculture alone makes a major contribution to river and canal pollution or whether, alternatively, aquaculture can place itself principally as the victim of pollution from urban and industrial sources [13].

A third concern about Thai aquaculture, alongside most agriculture systems globally, has been expressed: that emergence of AMR in environmental or animal-borne pathogens is driven by AMU on aquaculture farms, resulting in the increased potential for AMR infections in humans [14–17]. Indeed, the rapid growth and spread of the Thai marine aquaculture industry in the early 2000s was characterised by high profit margins sustained by a wide range of chemical inputs (including the use of antimicrobials) to prevent and treat diseases as well as improve water quality [18]. These inputs took place in an environment where many production-related diseases were present and unchecked, where increasingly concentrated fish and shrimp species were vulnerable to disease outbreaks and where, initially, a relatively unregulated legal framework governed AMU in animal farming [19–21]. The result was that much of the Thai aquaculture sector developed with relatively high levels of prophylactic AMU being the norm, particularly in the coastal zone [22].

Recent years have seen establishment of several significant regulatory and policy initiatives in Thailand to survey, monitor and reduce AMU in aquaculture [23,24]. Under these regulations, only the antibacterials oxytetracycline, tetracycline hydrochloride, sulfadimethoxine, trimethoprim, sulfadimethoxine/ormetoprim, amoxicillin and enrofloxacin are authorised in aquaculture [23]. Several market initiatives have followed suit with major retailers and food processing groups seeking to limit AMU within their Thai supply chains (e.g. the Raised Without Antibiotics initiative) [25]. The first five-year National Strategic Plan on AMR in Thailand was published in 2016, covering the period 2017 to 2021 [26]. Since then, legislation has tightened the availability and accessibility of antimicrobials for aquaculture and reinforced the role of the veterinarian in antimicrobial prescribing. Moreover, AMU for growth promotion has been prohibited, and certain antimicrobials considered critically important for human health have been made unavailable for use in aquaculture. These changes were in parallel with those in many other major international food-trading nations.

### Investigating current aquaculture practice

Fifty kilometres outside Bangkok, a wide river flows south through the agricultural district of ‘Plakat’ [a pseudonymised district name has been used to maintain anonymity], part of a larger administrative province. This is a flat fertile landscape with an elaborate network of irrigation systems and canals, bearing witness to agricultural and industrial activities following a period of land reformation. Rice fields and other field crops, vegetable farms, fish ponds, shrimp ponds, chicken, duck and pig farms are numerous in the district. There are also industrial factories and solar energy power stations. In agricultural terms, over the last two decades there has been a notable shift (as there has across much of central Thailand) from the more longstanding production of rice to more profitable, low salinity freshwater fish (notably tilapia) and shrimp production. Local fish markets serving Bangkok and nearby provinces provide critical commercial infrastructure, along with a range of aquaculture sector industries which include food-processing factories as well as feed and animal suppliers. With such animal production - and its associated commercial and market exigencies - come new and different pressures notably around health (both animal and public health), disease management and resource control, which interlink with the growing national agenda of antimicrobial surveillance and reduction mentioned above.

In an effort to identify the principal drivers of AMU in aquaculture and understand how inland, small-scale aquaculture farmers respond to shifts in animal health practice and regulation, our team spent a year in Plakat district, conducting research with freshwater prawn and fish farmers as part of a wider project to build a holistic picture of AMR drivers in Thailand from the One Health (human-animal-environment) perspective [27]. We investigated aquaculture farm management practices and attitudes towards AMU as well as the information and advisory networks they had for information about antimicrobials and aquatic animal health.

## Materials and Methods

A mixed methods approach was employed comprising a cross-sectional survey along with semi-structured interviews.

### Materials and participant recruitment

Primary research was undertaken through a questionnaire-based farmer survey supplemented by additional information obtained by a parallel series of semi-structured local community (household) interviews, undertaken as part of this research programme but oriented more specifically to human health and AMU (Fig 1).

**Fig 1.**
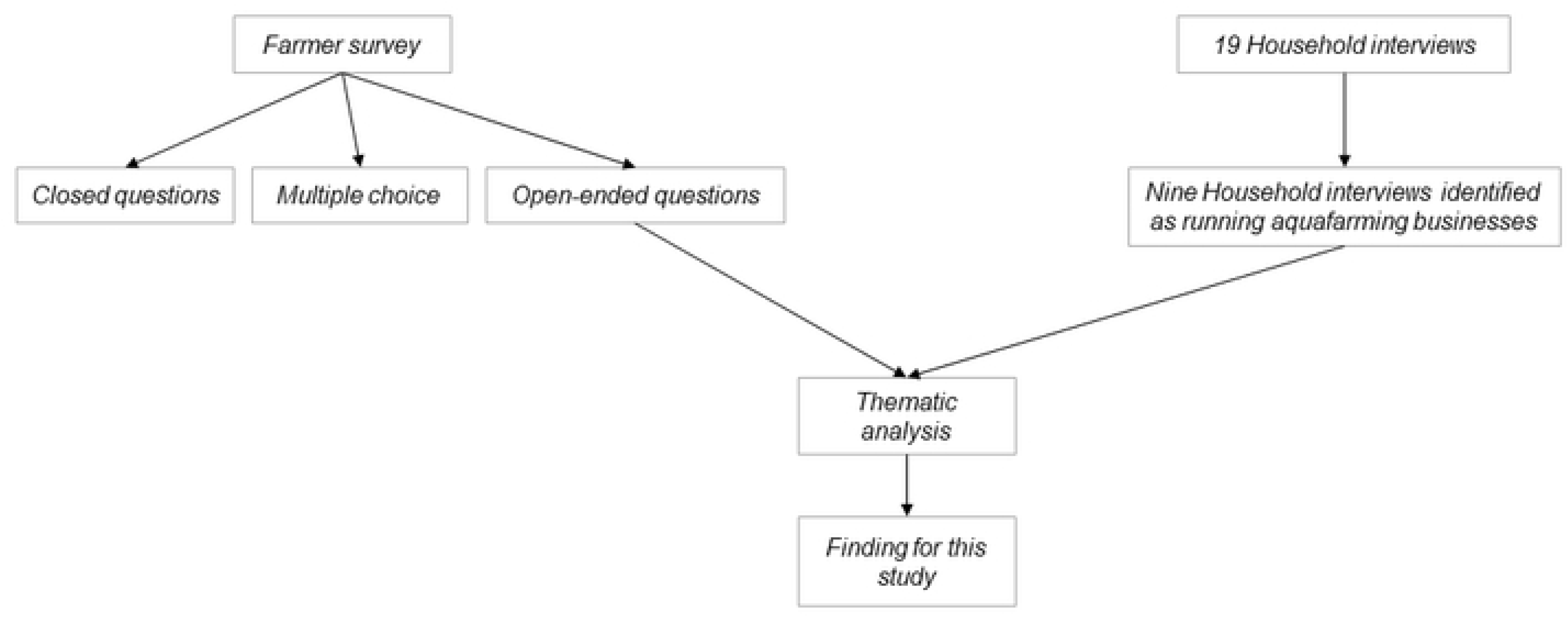
A diagram showing the methods and the data sources involved in this study.

#### Farmer survey

The purpose of the questionnaire survey was to explore the aquaculture farms and animal health management practices as well as AMU. A questionnaire was developed by the Thai and UK research teams that combined closed, open-ended and multiple choice questions covering three main areas: first, farm structure and farmer demographics; second, animal production and health; and third, medicine use including knowledge, advisory systems and networks used by farmers, with a particular emphasis on antimicrobials (Farmer survey in S1 Appendix). Data collection was conducted between October 1, 2019, and ended on December 20, 2019. A purposive sampling was followed based on geographical area and farm type. The farms were geographically centred around the principal settlement of ‘Plakat’ but stretched across the river basin and extensive local canal system. A total of 20 aquaculture farms were recruited: nine fish farms and 11 shrimp farms. The survey was conducted face-to-face (social distancing in place due to COVID-19 pandemic) with the farm owner by different members of the research team (BC, SS and/or AW). On a few occasions, co-workers were present and contributed to the discussion. Informed consent was obtained from all participants. All participants needed to sign a consent form before participating in the study. Only adults participated in the study. Open-ended questions were all in Thai, were audio recorded and subsequently translated to English and analysed. Responses to the closed and multiple-choice questions from the questionnaire were analysed and will be reported in future manuscripts. (BC, SS, AW) are researchers with a speciality and training in veterinary sciences.

#### Additional interviews with households

Additional qualitative interviews were undertaken with local residents, including aquaculture farmers, in the district as part of the wider project’s aim to understand local attitudes towards human health, AMU and AMR. These also provided additional information on aquafarming and animal health practices. Nineteen households were interviewed by researchers (KW, KP and NC) and later analysed for the aquafarming theme. Data collection took place between February 2019 and August 2021. The sample was purposively selected based on geographical area and socio-economic diversity. Initial contact was made by the village health volunteers (community health workers) who introduced the study on behalf of the research team [28]. Households who were happy to participate were introduced to the researchers. All visits were made on an invitation/permission basis from the key informant of the household. Social distancing rules were applied due to COVID-19 pandemic. On average, at least 5 visits were made to each household. Monthly visits were made to some households with which the research team had developed deeper relationships. Time per visit varied from the early (30-60 min) to the later (1-3 hours) period of fieldwork. An informed consent was obtained from all the households. Only adults participated in the study. Other adult family members contributed to interviews if they were present. Interviewees were in effect self-selected from among family members according to availability, but efforts were made to ensure a representative spread of occupations and ages across the overall sample. The three main themes for investigation included living conditions (livelihoods, food and water sources), health and treatment-seeking (including medicine use) (Interview topic guide in S2 Appendix). Researchers used both audio recorders and field notes to record these visits. Out of 19 households, almost half (nine) were identified as running small aquafarming businesses. (KW, KP, NC) are trained qualitative researchers with background in anthropology.

### Data management and analysis

Interview transcripts and responses to open-ended questions were recorded, transcribed and translated into English. These transcripts were then coded using open coding and analysed thematically [29] using NVivo qualitative data analysis software (Version 12, QSR International, Melbourne, Australia). A thematic analysis approach enables exploring the data in depth and identifying patterns of meaning across the data. The first step involved NA reviewing the transcripts through the stages of familiarization and assigning initial inductive codes to a sub-sample of transcripts. An initial coding framework was developed and discussed with the rest of the research team. The remaining transcripts were then indexed by NA using the coding framework and refinements were made as necessary. NA then re-read, reviewed and clustered the identified codes to form inductive themes and sub-themes and assigned names to the themes. The process was iterative and since the data were translated from Thai, cross-checking of accuracy of interpretation went on throughout the analysis through continuous discussion with the Thai researchers and the rest of the research team. Illustrative quotes from the farmer survey are presented with descriptors including the farm type (fish or shrimp) and the farm ID number (e.g. fish farmer F3 or shrimp farmer S4). Illustrative quotes from household data are presented with descriptors including the farm type (fish or shrimp) and household ID number (e.g. fish farmer HH38).

### Ethics

Formal ethical approval was received from Mahidol University Social Science Institutional Review Board [(Certificate of Approval No. 2019.026.0702), (MUSSIRB No. 2019/024 (B2)], and Faculty of Veterinary Science, Mahidol University – Institute Animal Care and Use Committee (FVS-MU-IACUC), (license no: UI-01292-2558) and registered with University of Bristol, UK.

The study used mixed methods including a survey of farmers and semi-structured interviews with adult members of households in the local community. For the farmer survey, written consent was obtained from all participants. For semi-structured interviews, written consent was requested before the commencing of data collection from all households. Some households initially preferred to give a verbal consent. In this case, the verbal consent was obtained and witnessed by another member of the research team, other household members and village health volunteers (Community health workers) who helped in the recruitment. Subsequent confirmation of consent was ensured on an ongoing basis (e.g. repeated requests for permission for household visits), in line with the Association of Social Anthropologists of the UK (ASA) Ethical Guidelines for good research practice (ASA 2021). Later, written consent was obtained from all households for other data collection activities (e.g. household survey, stool samples, reported elsewhere). For each activity, the research team explained to participants the study objectives and procedures, the voluntary nature of participation, principles of confidentiality and anonymisation of data. The consenting procedure (written or verbal) was approved by Mahidol University Social Science Institutional Review Board.

## Results

### Farm characteristics

Most farmers had members of their families living on the farm and working with animals. All farms were owned by the farmer; none were contracted from a company. Fifteen farmers had established the farms by themselves, three were inherited and two rented. Fish farmers mainly raised tilapia (*Oreochromis niloticus*). Other species included giant gourami (*Osphronemus goramy*), Jullien’s golden carp (*Probarbus jullieni*) and other carp species (Cyprinidae). Shrimp farmers raised Pacific white shrimp (*Litopenaeus vanname*i) and giant freshwater prawn (*Macrobrachium rosenbergii*) (Farm characteristics in S3 Table).

### Household characteristics

It was very common for household members to have multiple occupations (Household characteristics in S4 Table). Of the nine households, seven had private ponds used to raise fish or shrimp. Others had a cage (or floating baskets) used for raising fish in shared ponds or in the river (Fig 2).

**Fig 2.**
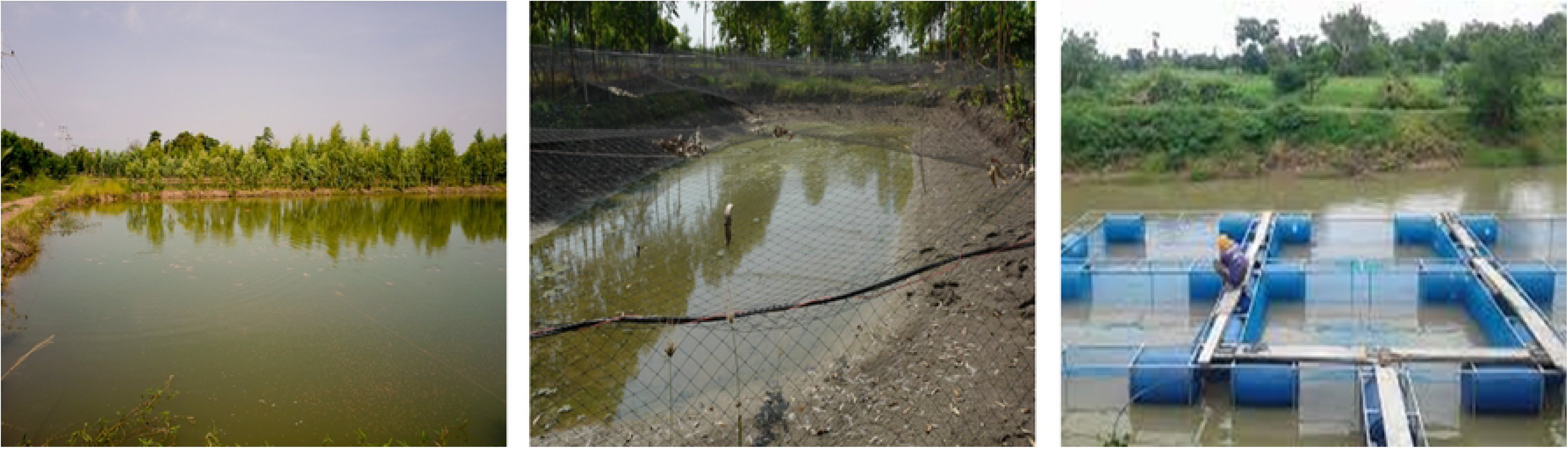
Left: Fishpond. Middle: Drained fishpond with protective nets. Right: Shared fishery in a pond or river.

Three main themes and nine subthemes were generated from the analysis (Fig 3).

**Fig 3.**
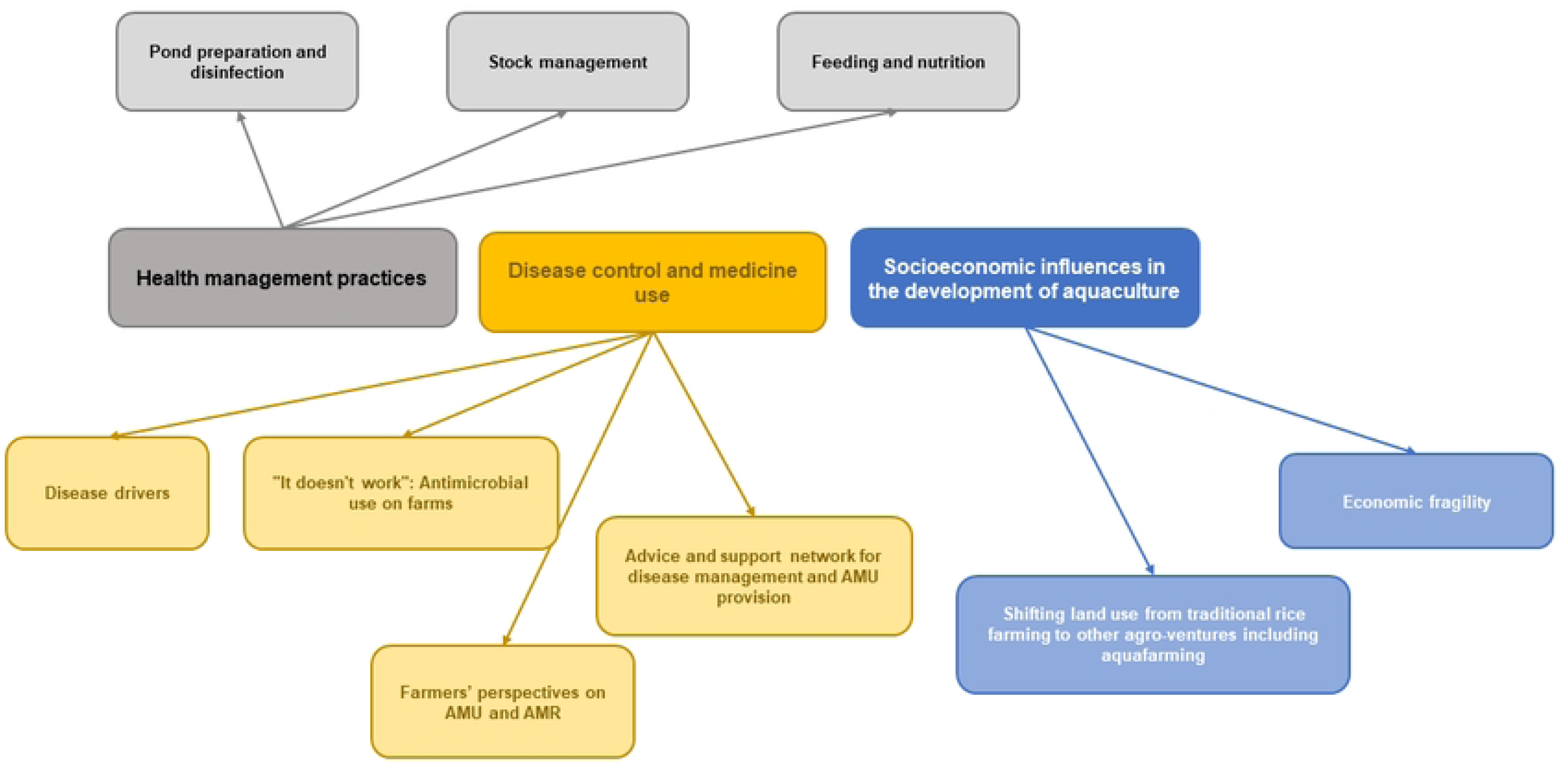
Thematic map of the themes and subthemes identified in the data.

### Health management practices

#### Pond preparation and disinfection

Farmers grew fish and shrimp in ponds dug out of the ground. To prepare the ponds for new stock, farmers would leave emptied ponds to dry out, clean the floor of the ponds and then add new water. Drying periods varied; one farmer spoke of allowing the floor to dry for 40-60 days while others mentioned a few days to two weeks. Once the soil was dry, most farmers scattered lime (calcium hydroxide or calcium carbonate) on the pond floor to disinfect the ground. Farmers also reported using other chemicals including herbicides or quicklime (calcium oxide) with two interviewees speaking of disinfecting their ponds with ‘germ-killing’ medicines (a local term generally used to refer to antibiotics or disinfectants). Treated ponds were left between one to 10 days, before water was added.

> *“We put in lime first to kill the germs…*
>
> *Researcher: For how many days, approximately?*
>
> *“Three days.”* Shrimp farmer S4
>
> *“To prepare the ponds, we fill them with water. Fill it up to the height that we think the fish can go in. Then disinfect the ponds using ‘germ-killing’ medicines.”*
>
> *Researcher: Do you also use lime before [putting the water in] at all?*
>
> *“No, no need.”* Fish farmer F8

Almost all farms obtained water for the ponds from the canal system connected to the river. Water levels were monitored and usually adjusted to a level of around one metre depth. Additives were often added to the ponds. These included salts, micro-organisms, ‘germ-killing’ medicines and fertilizers. The pond water then was usually left for around a week before the fish or shrimp were introduced. One farmer mentioned waiting for the water to ‘turn green’ before adding juvenile shrimp. Some farmers also checked the pH of the water.

> *“We scatter microbes [micro-organisms] and germ-killing medicines into the ponds, then we fill in water. After three days or a week, the shrimplets [baby shrimp] can go in.”* Shrimp farmer S5

> *“Sometimes we also add salt into some ponds to treat the soil and the water, to not let the water be too acidic or basic.”* Fish farmer F5

> *“[…] add water. followed by tablet fertilizer to prime the floor. That’s it.”* Shrimp farmer S10

> *“Interviewee: When the water goes in, we add microbes [micro-organisms] to treat it first*.
>
> *Researcher: That’s before you put the shrimps in*.
>
> *Interviewee: Yes. We use lime and microbes*.
>
> *Researcher: Do you buy the microbes or do you make that yourself?*
>
> *Interviewee: We buy. Sometimes we use [product name]. We use [product name] to ferment first, then we use [another product] to adjust the water condition…*
>
> *Researcher: From then, do you leave it for many more days?*
>
> *Interviewee: We leave it for another week before we put the shrimp in.”* Shrimp farmer S4

#### Stock management

Buying the aquatic animals from trusted sources was vital to farmers as such a practice meant fewer diseases later. New fish were usually kept together in a single, protective ‘nursery pond’ for an initial two and a half to three months. When the fish had grown in size and weight, they were divided between different ponds. Just over half the farmers reported co-housing different species. It was common for shrimp farmers to raise giant freshwater prawns and white shrimp together in the same pond. One farmer raised fish and shrimp together. Other animals like poultry (in small-scale production) roamed freely around the ponds or walked on the nets covering the ponds.

> *“But if you want to raise the fish together with the shrimp, you have to put the shrimp in first. We take that water to test in a lab; there’s a lab in [town]. If the water passes [the standards], then we can release the shrimps. Fifteen days after the shrimps go in, the fish can go in – tilapias and seven-striped carp and Chinese fish. Then we raise them.”* Fish farmer F7

#### Feeding and nutrition

Farmers explained that the aquaculture farming rearing cycle was generally operated over a one-year period. This period could be reduced to six to eight months depending on the farmer’s feeding style, whether they used organic feed or commercial supplements. Commercial supplements accelerated the growth of fish and shrimp, but not everyone could afford them. Some farmers used animal manure, sourced from their other livestock such as ducks, chicken, or pigs, to feed fish or shrimp in the ponds. This practice was seen as a way to reduce cost as well as being wise use of resources. One farmer mentioned having direct pipelines from their pig sheds through which dung was pumped into the ponds. Another farmer sold the extra dung they had to other farmers. Animal manure usage, however, was disapproved of by some other farmers due to its unpleasant smell. Farmers also reported using other types of organic or locally available sources of fertilizers such as vegetables from their garden, lotus, hay or rice bran either alone or mixed with powdered fishmeal usually given to small fish. One farmer mentioned using the wastewater from a fertiliser factory and another fed fermented pineapple peel to the shrimp. One participant described using unused noodles from a nearby factory. Vitamins were also used, though one farmer described them as a waste of money.

> *“I took pig dung to feed the fish. When I wash the pig dung from this coop, the water flows down two pipes; it flows into the fishpond.”* Fish farmer HH38

> *“Feed this fish only grain and rice bran. Mix rice bran with pellet food. Chicken manure, […] even the low cost, but the son doesn’t like it, he doesn’t want it. Chicken manure is smelly too. Big pond over there [ different farm]. The owner of the big pond raised it with chicken manure. Feed both pellet food and chicken manure. That fishpond was delivered by a truck of chicken manure. Feeding the fish like that makes the fish grow fast.”* Fish farmer HH03

> *“The noodles were brought to the fish to eat, but the noodles had to be marinated. Other farms do this. It has to add molasses water to mix in the tank as well. They buy from sugar factories.”* Fish farmer HH03

> *“I feed the fish with leftover food, vegetables that I bought to cook and then I throw the rest to the fish in the pond.”* Fish farmer HH25

### Disease control and medicine use

#### Disease drivers

Many farmers reported that diseases amongst their stock had increased over the last decade. While some farmers offered no explanation for this, others felt different factors were responsible including weather changes, the emergence of new diseases and decreasing water quality due to pollution.

> *“In the past they never got sick; if we found a few sick or dead ones we will just remove them.”* Shrimp farmer S11

> *“Lately, we don’t know what’s wrong with the fish, but they keep dying.”* Fish farmer F6

> *“If the water is good and the weather is good, we don’t need to use any medicine.”* Fish farmer F4

There was no consensus whether it was the heat or the cold that caused disease in the fish or shrimp, but it seemed that the extreme weather conditions led to disease spreading.

> *“The disease outbreak is usually at the end of winter/beginning [of] summer. Maybe, because of the heat, not sure, March-April. I give them medication and add a lot of water. I don’t know, maybe because it is getting warm around that time of the year.”* Fish farmer F6

> *“Researcher: You mentioned that the animals died a lot during the past year … how did it happen? What were the symptoms?*
>
> *Interviewee (1st male): It is the heat*.
>
> *Interviewee (2nd male): The weather is too hot, and they [the fish] got sick.”* Fish farmer and co-worker F7

> *“If the weather keeps getting colder, we will probably have to buy them medication.”* Fish farmer F9

> *“They are currently healthy, because right now the temperature is dropping. Once it got cooler, they are able to survive. But they would also start showing symptoms if it is getting too cold.”* Shrimp farmer S5

Water condition was considered vital in raising healthy aquatic animals and was monitored continuously, largely through attention to its colour, appearance and pH. Poor-looking water was often treated using additives such as salt, micro-organisms and ‘germ-killing’ medicines. Two farmers spoke of the impact of rain on the water conditions. Others spoke of the importance of oxygenating the water, sometimes artificially. Critically, these water quality and environmental factors played a part in determining subsequent antimicrobial and other medicine use.

> *“After one month, we add more water and salt. We have to take note of the water colour. If the colour starts to look bad – if it’s not green or clear – then we have to add salt or minerals.”* Shrimp farmer S9

> *“It depends on the weather and the water. For example, when we left them out during the rain, then they would start dying. They are unable to survive under that condition, because of the dropping pH, and alkaline, the shrimps are unable to adapt.”* Shrimp farmer S5

> *“I think the rain is toxic. I think. My opinion, not sure how true it is.”* Fish farmer F8

> *“It depends on the water. If the water quality is good, it would be acceptable for us not to add any medicine.”* Shrimp farmer S3

Half the farmers reported having significant animal disease outbreaks or mass dying events recently, with six reporting estimated mortality rates of over 20% - in one case, rising to 80% of stock. One farmer described disease symptoms in fish as *“red belly, disease in the eye, swollen navel”* (Fish farmer F9). Another looked for specific symptoms such as bruises as signs of disease. For most, shrimp or fish feeding rates and mortality levels served as the basic indicators of animal population health. One farmer mentioned using a lifting net to examine the fish closely or shining a light at them at night.

> *“The amount of death is uncertain because we can see only when the dead fish are floating.”* Shrimp farmer S2

> *“When they got sick, when they started to die. Once they died, they will float, and we just need to determine what kills them. Like, if they have bruises, we would have a look and decide on bacterial or viral, and then we would treat them accordingly.”* Fish Farmer F6

> *“Researcher: From your experience, you lift Yo (a tool for trapping fishes) to see how they are?*

> *“Yes. We also can see when the fishes come up to the side of the bank. We shine the light to see them during the night time… we check whether they (shrimp) have tight texture or black gum…have foods in their intestines.”* Shrimp farmer S7

#### *“It doesn’t work”*: AMU on aquaculture farms

Farmers were generally divided as to whether AMU on aquaculture farms had increased or decreased in the last decade. Those that mentioned AMU decreasing argued that medicine cost was high and that aquaculture farming simply did not generate enough revenue to cover this cost. Some doubted the benefits of using antimicrobials in treating aquatic stock, believing antimicrobials actively stunted growth. For a few respondents, government regulations along with food chain and market imperatives mitigated against the use of antimicrobials.

> *“[AMU] decreased, to decrease the running cost of the farm… Because, antibiotics are expensive and sometimes ineffective.”* Shrimp farmer S9

> *“Mostly, they [aquaculture farmers in general] don’t use [antimicrobials] anymore… It doesn’t work.”* Fish farmer F4

> *“However, even medicines did not really help… when we give them medications it looks like they won’t grow, it looks like the medicines stunted their growth.”* Fish farmer F7

> *“I heard that people say it’s not good if we use too much. If we sell the shrimps and the merchant/buyer checks and detects the drug use, we won’t be able to sell our shrimps anymore. That is the reason why I try to limit the usage of medicine with my shrimps.”* Shrimp farmer S3

> *“For medicine, we may not use it at all because the Department of Fisheries already told that it will be problem if medicine is detected. We used to face that problem.”* Shrimp farmer S6

Such farmers preferred to use other methods to control disease. These included selecting reputed aquatic animal providers from which to purchase stock, using herbal medicines and adding vitamins to enhance growth. As stated above, water quality was seen as an important factor in keeping the stock healthy, often with additions of salts and lime. In the case of major disease outbreaks, farmers regularly removed any floating dead fish and shrimp and would deploy emergency harvesting if needed. Emergency harvests were either sold for sauce production or became landfill. Infected ponds were then disinfected, and water was pumped out to prepare for a new batch.

> *“Mostly, I don’t use [antimicrobials]. Because we focus on the species (of aquatic animals) that we got from different farms…they are not similar.”* Shrimp farmer S6

> *“Now people mostly use organic methods… we just won’t use it [antimicrobials]. We will concentrate on natural/organic ways.”* Shrimp farmer S4

> *“Yes. I use only herbs I bought from the food shop. I remember that it made from turmeric, which human also can eat … If vitamins, we use sometimes … When I find that some ponds grow slowly, I make a decision by myself to use vitamin supplements.”* Shrimp farmer S6

> *“To avoid using antimicrobials, we prepare the pool and water by adding microorganisms; oxygenate the water, to help strengthen the animals, so that they won’t be bothered by any diseases.”* Fish farmer F3

> *“Researcher: What did you do with the dead ones, did you bury them? Or sell them? Interviewee: We sold them. We sold them as “Phla-ra” [salt-fermented fish (not fish sauce) commonly used in many Thai dishes]. Thirteen tons of Phla-ra.”* Fish farmer F8

> *“During [disease outbreak] we catch them; if we see they die, we must hurry to catch them. If we let it be, they will all die.”* Shrimp farmer S8

Other farms surveyed reported that their use of antimicrobials had increased in recent years in response to growing disease prevalence. Prophylactic use of antimicrobials was often described, particularly when an outbreak on a neighbouring farm was discussed. As a treatment, antimicrobials were largely considered to be a last resort and were far from being seen as being effective in treating sick animals or populations. Most farmers were unable to name specific antimicrobial medicines, recognising them rather by their appearance or characteristics.

> *“I think the use [of antimicrobials] is increased. Nowadays, diseases of aquatic animals are more and more every day. We need to use antibiotics for curing their diseases.”* Fish farmer F2

> *“However, I occasionally give them medicine for disease prevention if there is a disease outbreak and others say I should give them as a prevention measure.”* Shrimp farmer S4

> *“[…giving them antimicrobials] To stimulate them. […] To prevent them from getting sick… we don’t give them that often, sometimes we would refrain from giving them the medication. If they look responsive, we won’t give them any.”* Shrimp farmer S3

> *“We don’t know what else we could do. We already try changing the water, and aerating; we already did everything, but it didn’t work. So, we have to turn to using medications.”* Fish farmer F6

> *“[…] Like when they ate [the medicine] for seven days in a row, they stop [dying]. However, if we stop giving them the meds, they will start dying again. They are not completely cured. We don’t know how to solve this.”* Fish farmer F8

> *“If we see giant freshwater prawns die, then we use yellow medicine. That’s it. Nothing much here because we don’t want to invest too much. The price of the shrimp [to sell] is low.”* Shrimp farmer S3

Out of the 20 farms, only six farms reported using antimicrobials including oxytetracycline, fluoroquinolones (enrofloxacin, norfloxacin and ciprofloxacin) and sulphonamides. Farmers reported purchasing antimicrobials from feed companies or stores and mixing these with feed on the farms. Farmers did not mention obtaining antimicrobials from veterinarians. Other additives were also used including multivitamins, amino acids and probiotics. The dosage and frequency were mostly determined by the farmer and varied significantly between farms, suggesting the absence or lack of knowledge of common guidelines within this small-scale commercial aquaculture sector. Treatment plans were pond- and population-based. When some fish or shrimp showed signs of sickness, the entire pond would be treated with the medicines mixed with the feed or water and spread manually into the pond.

> *“There is a powder medicine that we can mix in water and put in the pond.”*

> *Researcher: That would be for the entire pond?*

> *“Yes. Some people would mix it in the food. It’s different for each farm.”* Shrimp farmer S11

> *“We cure them all in a pond. If this pond was cured, I can leave them and take care of the next pond.”* Shrimp farmer S8

#### Farmers’ perspectives on AMU and AMR

Farmers were asked what they thought about the long-term impact of AMU on humans, animals or the environment. Misinformation or lack of awareness was evident as some farmers felt there was no negative impact from AMU.

> *“I haven’t seen any effects from the use of antimicrobial so far, neither has anyone come to clarify/educate us about it.”* Fish farmer F3

> *“It’s not harmful to humans… My opinion would be that our use of medicines isn’t harmful and doesn’t affect the consumer; even the biologist that gave us the medication said that there is no effect.”*

> *Researcher: What about these medicines – germ killer? Do you think they have any effects on the consumer?*

> *“No.”* Shrimp farmer S4

> *“Researcher: Do you know about the effects of using antibacterial drugs on human, animals and environment? Is there any effect when we use them for a long time?*

> *“Interviewee: No, I don’t know. I don’t really study about this, so I don’t know much.”* Shrimp farmer S7

Other farmers believed that AMU could affect consumers due to chemicals accumulating in the body and subsequently causing illnesses. However, these farmers explained that using antimicrobials was necessary, especially in large farms with many animals.

> *“I know that if we use a significant amount of medicines with the animal, it will affect the consumer as well. The antibiotics we used would accumulate in the human body, causing it to resist drug action afterwards. So, yes. I know that it would cause drug resistance in the human body when we use it to treat illness later on.”* Fish farmer F6

> *“If we use too much medicine for a long time, it would definitely be harmful to humans because it would accumulate in our body. But it is necessary to use medication because we need to medicate the animal when they get sick, especially for those big farms that have a lot of animals.”* Shrimp farmer S11

#### Advice and support networks for disease management and AMU provision

The farmers interviewed stated that they had autonomy over the health management and treatment decisions taken on their farms. Farmers tended to learn what treatment to use through their own experimentation with different products and their impact upon illness symptoms and animal health.

> *“I decide from the symptom whether to use Oxy [oxytetracycline], Sulfa [sulfonamides], Enro [enrofloxacin], or others. There are 2 to 3 primary medicines that I use nowadays.”* Fish farmer F8

> *“If I use any product, and it’s good, then I continue using that [laugh]… I have no one to do research for me. So, I must do it myself. If I give the fishes any medicines and see that during the three days they get better, then I continue using that one*. *But if they don’t get better within three days, then I have to change the medicine.”* Fish farmer F8

Some sought treatment recommendation and advice from more experienced peers, animal husbandry personnel and from storekeepers who sold aquatic food and medicines. Farmers occasionally referred to these sellers and personnel as ‘vets’; farmers would describe the symptoms to the sellers and the sellers dispensed the medicines. Some stores also offered water testing services.

> *“I asked the store owner too, and other people who used the same products or had the same experiences. So, they can give me advice on what to do or what medicines to use.”* Fish farmer F3

> *“I would consult with people at the drug store. They usually have a veterinarian there. Sometimes I would ask the veterinarians who can prescribe medicine that I know. I would tell them the symptoms - such as having red scales, red gill and droopy eyes - then ask which medicine I should use. They would give suggestions based on that. We don’t meet face-to-face for the veterinarian to check the water condition and tell us exactly which medicine we need to use specifically for such a disease. I would be more sure like 100% if that is the case.”* Fish farmer F8

Dedicated aquatic veterinary support was considered very rare in the area and farmers took every opportunity to contact a veterinarian whenever one came into the neighbourhood, though none of the farmers interviewed reported a veterinarian visiting their farms for routine checks or in case of an illness or outbreak. One farmer said he had attended a training course on aquatic farming, and another said they had done their own reading.

> *“At the beginning, I usually ask the agriculturist/academic. But after a while, I started to study things myself. I tried different things while seeing what other people use. That’s how I ended up with this product. Actually, others suggest me to use Amoxy [amoxycillin] as well.”* Fish farmer F6

> *“A fisheries scientist used to come to the district office, so I memorized the names of the medications. I would call and asked him, whether this meds would work or not, and he said that it can. So, I made the decision.”* Fish farmer F6

> *“I attended training about fish diseases before. The instructor instructed us to use which medicine with what symptom. I had the instructor’s phone number, so I used to call him and ask him for the suggestion of medication relating to the fishes’ symptom, but I lost the number.”* Fish farmer F6

### Socioecological influences in the development of aquaculture

#### Shifting land use from traditional rice farming to other agro-ventures including aquafarming

Many participants described how land was, in the past, mainly used for rice farming and that rice fields spread as far as the eye could see. Participants mentioned that, during the 1990s, the use of land started to shift due to a fall in rice prices and subsequent economic loss. Additionally, participants explained that the rice farming workforce has reduced over the years due to aging with younger generations not wanting to undertake low-income occupations. Subsequently, new business ventures started to appear. Factories were built and other agricultural systems were promoted including gardening, plantations, aquaculture farming and livestock farming to generate more income.

> *“Many people are doing the shrimp pond business because the income is good. Those who cannot do the rice field will have shrimp ponds. The shrimp pond business has been around for about 10 years because people had left the area and when they came back, they started a shrimp pond business.”* Fish farmer HH25

> *“Our family switched to raising fish in the old fields, digging old fields into fish ponds. At that time, farming was not good. Oh, now farming requires a lot of investment, a lot of things. My parents are old and I can’t farm myself. So our family has switched to raising fish instead.”* Fish farmer HH25

> *“My family used to farm rice. I don’t plant rice anymore, now I raise shrimp. In the past, most of the rice fields were cultivated in this area, 80-90% of the area was rice fields. Today it is transformed into a pond and mixed farming. It has changed in the past 10 years from rice fields to shrimp ponds. However, when shrimp prices are not good, there are farmers who turn back to rice farming. But my family has always been raising shrimp since we have been raising shrimp and never changed. I’ve raised white shrimp before. I planted Manila tamarind on the edge of the shrimp pond, cultivated as an additional income. One year it can sell for over a hundred thousand baht.”* Shrimp farmer HH19

#### Economic fragility

All the households involved in aquaculture farming also had other sources of income. Household members worked in various agriculture projects such as rice farming, growing vegetables, fruit gardens, lotus ponds and aquaculture farming as well as livestock farming including poultry and pigs. Other occupations such as working in factories and cookery were also common. Participants explained that having different occupations provided more financial stability to support families. For example, one household had to close their chicken farm in 2006 due to avian influenza. However, they had other sources of income from raising shrimp and growing rice as well as working as a truck driver. Another household, which had a chicken farm contracted to sell eggs to a larger farm, closed the chicken egg business due to lower egg prices being imposed by the larger farm. They had to sell all the chickens back to the larger farm (from which they were originally purchased) at a financial loss. This household, however, also had a tilapia pond they could rely on. One participant explained that the concept of holding several farming projects was encouraged by the government-promoted “sufficient economy” ideology which advocated the wise use of one’s own resources and the redeployment rather than wasting of these resources.

> *“The people choose to do many kinds of agriculture because they might face a lack of income if one of their businesses has some complications.”* Shrimp farmer HH19

> *“My job is to raise fish and plant lotuses. My husband [HH227(02)] is the one who picked up the lotus from the pond. I also help my husband. My other job is raising my grandchildren… my husband will go out to feed the fish at 6 am. The pond has tilapia and yisok [Julien’s golden carp]. Our ponds often sell fish to fishmongers who sell fish in the fish market. They will come and catch fish at the pond themselves.”* Fish farmer HH27

> *“I repaired the equipment myself and also repaired the machines for other people in the community.”* Fish farmer HH27

> *“My house is sectioned according to the sufficiency economy’s prototype that is the two rai areas [about 3800-6400 square meters] of the land is for the house, various kinds of vegetable gardens, a fishpond and a pig shed. The rest of the land, about 36 rai, is paddy field which is shared with other types of agriculture.”* Fish farmer HH38

Notably, most households were multi-generation residencies where grandparents, siblings and grandchildren lived together in one house or adjacent houses on the same piece of land. This concentration of family members provided labour for these multiple occupations. Participants described helping each other in running different projects with stay-at-home members (elderly, unemployed) generally helping with commercial cooking (restaurants, catering) and aquaculture farming or livestock feeding.

> *“This fishpond belongs to my son. I couldn’t do it, I let the kids feed the fish. When catching fish for sale, there will be many relatives who are selling fish in the fish market… We don’t have to do anything. My son works at the university. He returned on Saturday and Sunday. My daughter is a fish feeder… This pond is raising tilapia, carp and yisok. It has been raised for 7-8 months.”* Fish farmer HH03

## Discussion

This study aimed to investigate the socio-economic and ecological conditions of freshwater aquaculture farming and the drivers for AMU in a district in central Thailand. Our findings demonstrated that there was limited use of antimicrobials across the farmers in this study. Those farmers expressed doubt about the benefits of antimicrobials, both in relation to disease treatment and aquatic animal growth. High cost and prohibitive market regulations also discouraged farmers from using antimicrobials. Disease drivers were often linked to weather changes, the emergence of new diseases and decreasing water quality due to pollution. Farmers depended on improved farm management practices to maintain the health of the aquatic animals; when that failed, many farmers applied emergency harvesting techniques (i.e. no treatment but catching animals that were still alive). Aquaculture veterinary support was limited in the area and many farmers relied on their social networks and on experimenting with medicines in relation to decision making about disease treatment. Aquaculture farms in this area took the shape of domestic ventures operated by family members. These families were often found to invest in different juxtaposed small businesses in an attempt to maintain or improve the financial stability of the household. Aquaculture farming was considered a profitable business that did not require high input and labour. Initial outlay was often reduced by repurposing unused crops, food or animal manure to fertilise the ponds and stimulate aquatic animal growth instead of, or in addition to, using commercial feed.

It is interesting that many farmers in this study stated that they did not use antimicrobials in their farms. Farmers referred to antimicrobial treatments as ineffective and expensive. Traditional antimicrobial administration methods are often problematic and complex in aquaculture; dispensing antimicrobials directly into water or adding antimicrobials to aquatic feed can be ineffective, possibly harmful and could lead to AMR [30–33]. This is due to difficulties in adjusting a therapeutic dose uniformly across the pond, the effects of the natural environment on the antimicrobial as well as associated toxicity (e.g. to denitrifying bacteria leading to a build-up of toxic ammonia) [31]. Additionally, diseased fish often do not eat, which reduces the success rate of medicating the feed [31]. Accordingly, specialist advice is important to facilitate effective antimicrobial administration in aquaculture systems. It is possible, therefore, that, in our study area, due to the lack of veterinary support, our participants’ own attempts at antimicrobial treatment failed. This may have resulted in their opinions about the lack of effectiveness of these drugs, which led them to stop using antimicrobials and look for alternative disease management options. However, a few participants still reported using antimicrobials despite limited observed benefit. Other studies have noted that antimicrobial use may fluctuate from year to year even in the same area, subject to changing climate and disease incidence; one review article noted that farmer surveys often described inconsistent reports of antimicrobial use [34]. Another study in Vietnam that monitored antimicrobial use found that although 45% of farmers believed antimicrobials had no effect on curing diseases, 86% of the farmers who held that opinion still used antibacterials for either treatment or prophylaxis [35]. These reports suggest that our results indicating low use of antimicrobials should be interpreted with caution; although the findings indicate little use, it would be valuable in our study area to see whether abstinence from AMU continues into the future. Another relevant issue to AMR is the reported use of fluoroquinolones in this study. One attraction of using fluoroquinolones in aquaculture is that these molecules are stable in aquatic environments, rendering them easy to manage for aquaculture disease treatment [36]. Indeed, enrofloxacin is widely used globally [24]. However, the use of fluoroquinolones in aquaculture has been banned in many countries due to their importance of some fluoroquinolones to human medicine (especially ciprofloxacin and norfloxacin). This is because when resistance to one fluoroquinolone (whether used only in farming, or used to treat animals and humans) emerges, it usually confers resistance to all fluoroquinolones [15,24].

Limited AMU in aquaculture, as was observed in this study, would broadly be considered positive in relation to AMR; to ensure its sustainability, however, improving farm management practice is also vital to compensate for the health needs of the aquatic animals as well as to ensure farm biosecurity. Farmers in this study adopted several management practices they considered important to protect their stock from diseases: pond preparation and disinfection, healthy stock acquisition and good nutrition regimes, to name a few. These farms resembled the description of the small-scale aquaculture farms described by the work of Little et al. [10,37] which describes aquaculture as an activity practiced predominantly by farmers “for whom aquaculture constitutes one element of a larger total livelihood portfolio”. In these systems, farmers often adopt lower-input and lower-risk practices similar to extensive farming systems. This is, for instance, by crop diversification, improving pond preparation, choosing better-quality stock and using improved quality feed regimes, subsequently reducing disease incidence and the need for treatment [38,39]. Similar multi-culture agricultural systems such as home garden systems in Vietnam, Indonesia and Sri Lanka have been encouraged for their socio-economic and ecological benefits, and have been highlighted for their potential role in alleviating poverty [40–43]. In Thailand, the effects of the extensification and the diversification of crops, as seen in our study area, has been shown to lead to resilience in facing diseases (e.g. rotating rice and shrimp production showed reduced disease susceptibility when compared to back-to-back shrimp farming) [13]. Governmental support has also provided knowledge and production management plans which have, in turn, encouraged farmers to establish mixed cropping agriculture [18]. Land has often been divided into an area for the main farming activity, the secondary activity and a supplementary activity, and has included plans for production of different types of plants, livestock and fisheries [18]. Repurposing local resources such as we saw in in our study area (e.g. using left-over crops and food to feed aquatic animals) has also been encouraged. Another characteristic of Thai aquaculture farming was that farmers were seen to be well connected to vertical and horizontal knowledge networks [44]. Their vertical network went along the supply chain and also included government representatives, pharmaceutical and feed companies. Farmers also belonged to social networks of interlinked farmer groups and small clubs known locally as “Chum Rom”. These networks created a rural seminar culture where farmers shared their knowledge and practices, and industry and government representatives attended and added to discussions.

Despite improved management practices, findings of our study indicated that farmers were still facing high disease and mortality rates among their aquatic animals. This could risk the successful adaptation of extensive farming styles with low AMU, resulting in farmers switching to more intensified farming practices with higher AMU [36,45]. Reasons for high disease incidence were unclear but farmers cited water pollution as one of the drivers of disease occurrence. The effects of water pollution on aquaculture farming have previously been investigated in Thailand costal aquaculture regions [46,47]. These studies found that aquaculture water quality was indeed affected by urban pollution in the canals and included faecal coliforms, human *Escherichia coli*, tetracycline resistance genes and nitrogen [46]. The aquaculture water was a source of salinity and herbicides. Additionally, high AMR prevalence was found to be associated with a high prevalence of faecal indicator bacteria which was highest in peri-urban canal water feeding the aquaculture systems [47]. Another study in Taiwan investigated the potential cross-contamination problems between aquaculture systems and surrounding waters [48]. This work demonstrated that aquaculture activities (i.e. usage of antimicrobials) impacted the surrounding aquatic environments and, at the same time, the surrounding anthropogenic activities impacted aquaculture waters. These studies show that aquaculture (even with low AMU) would still be in danger of transmission of AMR from the environment. Another factor potentially contributing to environmental AMR transmission and selection in aquaculture systems is the use of animal manure in fertilising the ponds [34,36], a practice applied by some of the farmers in our study. A few of these even had integrated systems where manure was pumped from other animal sheds (e.g. pig or chicken) to the ponds. This practice was seen by farmers as economically sustainable. However, it is well established that manure is a reservoir of resistant bacteria and antimicrobial compounds [49]. For example, in a study investigating the impact of integrated fish farming (a practice combining livestock and fish farming, where animal manure is shed directly into fish ponds) on AMR, the level of resistance in *Acinetobacter* spp. was found to increase from 1-5% of bacterial isolates resistant prior to integration to 100% resistance to oxytetracycline and sulfamethoxazole and to more than 80% resistance to ciprofloxacin after two months [50]. Another study has shown the correlation between diffusion of fluoroquinolone resistance genes and biofertilizers utilisation in Chinese shrimp aquaculture [51]. There is certainly a need to raising awareness among aquaculture farmers of the risks of using contaminated manure as well as the wider role of veterinary and husbandry support.

Another relevant finding linked to drivers of AMU was that participants linked disease incidence to changes in climate, and particularly the effect of heat on fish health. Prophylactic AMU was seen more commonly when farmers anticipated weather changes. Interestingly, increased local temperature has been associated with increasing AMR in human infections and this association was consistent across most classes of antibacterials and pathogen [52]. Another study has found that increased temperature was associated with increased odds that faecal samples from the environment were positive for resistant *E. coli* [53]. If AMR were also to increase on fish farms as a function of temperature, it would be expected that antimicrobials would then have reduced ability to cure bacterial infections in treated animals. Increased temperature also affects the chemical activity and uptake of medicines in the pond environment [30]. All these factors mean a possible increase in disease incidence on aquaculture farms as the temperature rises, along with forward pressure on AMU, further exacerbating AMR. There is a specific need, therefore, to support farmers in developing disease management plans as the climate warms.

This study was a qualitative investigation, and the findings are specific to the research geographical area and may not be generalisable to other geographical areas. Future studies could include several areas with freshwater aquaculture. Farmers accounts of their use of antimicrobials, disease incidence and mortality rates are subject to recall bias.

## Conclusions

Farmers in this study reported limited AMU in small-scale family aquaculture farming due to intentional and unintentional socio-economic and ecological factors. Governmental support to encourage reduced disease risk and crop diversification as well as market regulation concerning the residues of antimicrobials allowed in food may have encouraged farmers to reduce their use of antimicrobials. Past experience of ineffective treatment of farmed aquatic animals with antimicrobials and the high cost of these medicines have also played a part in the low AMU reported. Increased disease rates were attributed to weather changes, the emergence of new diseases and decreasing water quality due to pollution. The lack of specialist aquatic veterinary support might leave farmers subject to pressure from commercial drug sellers when they seek disease management advice and misuse may also make treatment failure more likely, particularly as temperatures rise. There is a need to investigate the effects of climate change on aquaculture farming across Southeast Asia as well as its associated disease and treatments patterns. Further attention is needed to understand and raise awareness about the risks of using contaminated animal manure in aquaculture farming. Future policies should attempt to fill the gap in specialist veterinary provision to freshwater aquaculture, address the need to provide evidence-based information and advice on farm management practices that reduce the need for prophylactic and therapeutic AMU, and particularly address disease prevention in the face of a changing climate.

## Data Availability

Data cannot be shared publicly because of risk of participants identification. Data contains potentially identifying and sensitive information. Data will be available from University of Bristol Research Data Repository (https://data.bris.ac.uk/data/) for researchers who meet the criteria for access to confidential data.

## Acknowledgements

We thank all the community members and the farmers who participated in this study.

---

**S1 Appendix. Farmer survey**

**S2 Appendix. Interview topic guide**

**S3 Table. Farm characteristics**

**S4 Table. Household characteristics**

